# Secondary Vulvodynia Patients Show Reduced Bacterial Diversity in Vestibular Microbiome

**DOI:** 10.1101/2020.02.11.20021014

**Authors:** Anu Aalto, Pashupati Mishra, Sari Tuomisto, Tiina Ceder, Kati Sundström, Riikka Leppänen, Katja Ahinko, Johanna Mäenpää, Terho Lehtimäki, Pekka J Karhunen, Synnöve Staff

## Abstract

**Introduction:** Localized Provoked Vulvodynia (LPV) is a quite common form of sexual pain among young women. Primary LPV (e.g. pain from the first attempted intercourse) and secondary LPV (pain-free time period before the initiation of symptoms) have been suggested to develop via different pathomechanisms. As inflammation is one possible trigger in LPV, changes in vaginal and/or vulvar microbiome may be an initiator of the inflammatory process leading to peripheral neurosensitization and predisposing women to LPV.

**Aim:** Aim of this pilot study was to find out, whether there are differences in vestibular microbiome between LPV patients and controls.

**Materials and Methods:** Thirty women with LPV (8 with primary and 22 with secondary LPV) and 21 controls were prospectively recruited to the study from Kanta-Häme Central Hospital and Tampere University Hospital (TAUH). Paired vestibular samples were collected in clinical outpatient setting and analyzed with 16s rRNA sequencing.

**Results:** LPV patients had lower BMI, were younger and more often nulliparous than controls. Patients with secondary vulvodynia had reduced (p = 0.06) bacterial species diversity (alpha diversity) and more clustered (p= 0.05) vaginal microbiome community (beta diversity) compared to healthy controls. Differential abundance analysis identified 31 bacterial taxa that contributed to the difference in bacterial composition between secondary cases and controls (p < 0.05). Bacteria taxa that were increased among patients were *Gardneralla vaginalis* (p<0.0001), *Peptoniphilus* spp (p<0.0001), *Prevotella amnii* (p<0.0001) and *Streptococcus* spp (p=0.0001). Several bacterial species were more abundant among healthy controls reflecting reduced bacterial diversity among patients.

**Conclusion:** In patients with secondary LPV changes in vulvar microbiome may contribute to pathogenesis of LPV and ultimately lead to targeted therapeutic options. These preliminary findings need to be confirmed in a larger study.

## Introduction

Localized provoked vulvodynia (LPV) is the most common form of sexual pain among young women ^1^ and it is estimated that 8-15 % of women suffer from vulvodynia currently ^2–4^. LPV is described by burning, sharp or cutting pain in the vestibular area in response to different activities like penetration, tampon insertion, that cause pressure to vaginal opening^5^. The etiology of LPV is unknown but inflammation ^6^ and preceding genital infections such as yeast infection ^7^ are suspected to play a role in the ethiopathogenesis of LPV ^8^. Primary LPV and secondary LPV can be distinguished by the timing of first pain symptoms; in primary LPV, pain is present from the first attempted intercourse, tampon insertion etc.^5,9^. A pain free time-period before developing symptoms typically precedes secondary LPV.

The microbiome has been shown to affect human physiology, nutrition and immunity (Reviewed by Smith & Ravel, 2017). The microbiome of the vagina is under constant change caused by e.g. puberty and menopause and associated changes in hormonal status ^10^. Antimicrobial medication, sexual activity and menses affect the vaginal microbiome ^11^. The Human Microbiome Project found, using 16S rRNA sequencing, that Lactobacillus genus was dominant in vaginal microbiome at the vaginal introitus, midpoint and posterior fornix ^12^. The same project also found that vaginal microbiome is usually dominated by one or two Lactobacillus species that are most frequently Lactobacillus iners, L. crispatus, L. gasseri or L.jensenii which can fluctuate on the basis of the women’s menstrual cycle ^12^. Lactobacillus helps to keep the acidic environment in the vagina and prevents growth of yeast, harmful bacteria and viruses ^13^. Certain types of lactobacilli are harmful, if present, and at the same time the lack of dominant and healthy lactobacillus in vagina can predispose women to sexually transmitted disease (STD), bacterial vaginosis, pelvic inflammatory disease and preterm birth ^13^. The microbiome of the vulvar vestibulum is much less studied compared to the vaginal flora. However, the dominant bacteria of the vagina are mostly similar to the microbiome found in vulvar vestibulum ^14^ because vaginal secretions are an important source of microbial flora found in the vestibulum^14^. The alteration in the vaginal flora might be the initiation of an inflammatory process that finally may result in the abnormal cytokine production and development of LPV symptoms ^15^.

There is very limited data available concerning metagenomics between vulvodynia patients and controls in terms of vaginal or vestibular microbiome using 16S rRNA sequencing ^14^. This pilot study comparing the microbiome between LPV patients and controls was undertaken in order to evaluate potential differences that are possibly related to vulvodynia symptoms, with the ultimate aim to develop targeted therapies.

## Materials and methods

### Study subjects

Thirty women over 18 years of age suffering from LPV were prospectively recruited from the gynecological outpatient clinic in Kanta-Häme Central Hospital and Tampere University Hospital (TAUH). All study participants had to fulfill Friedrich’s two criteria ^16^ : The presence of pain on pressure to the vestibule or when attempting to insert an object into the vagina and pain on pressure to the vestibule upon examination. Symptom duration >6 months was required in the study. Exclusion criteria were: pregnancy, other current benign or malignant condition of vulvar vestibulum or vagina, under 18 years of age. Other causes of vulvar pain were excluded by careful patient history, and clinical examination to exclude herpetic lesions, trauma, fissures and clinical evidence of current infectious disease of the vagina and vulva. As controls we prospectively recruited 21 otherwise healthy women aged over 18 years who were admitted to hysteroscopy for a benign reason (e.g. polyp, anomaly of the uterus, bleeding disturbances) in Kanta-Häme Central Hospital or TAUH. The hysteroscopy procedure was done under routine general anesthesia or in ambulatory setting without general anesthesia. The vulvar skin was examined carefully and only patients with clinically normal vulvar skin were recruited. The exclusion criteria were: pregnancy related conditions (e.g. residua of abortion), other current benign or malignant condition of vulvar vestibulum or vagina, under 18 years of age. All patients and controls gave written informed consent before participating in this study. The study complies with the Declaration of Helsinki. Study protocol was reviewed and approved by Pirkanmaa Hospital District Ethics Committee (Identification code: R17081).

### Study samples

Vestibular samples were collected with buccal swabs (lFLOQSwab®, Copan Diagnostics, Italy) by rubbing two swabs against the vaginal vestibule (Figure 1). Samples were placed in Copan Diagnostics ESwab sample collection tubes (Fisher Healthcare, Houston, Tx) and stored at −70 Celsius degrees and shipped by express mail on dry ice to the Fimlab Laboratories Ltd., and stored there until further analysis at the laboratory of the Faculty of Medicine and Health Technology. DNA extraction from the samples was done with Macherey-Nagel NucleoSpin Tissue DNA extraction-kit (Macherey-Nagel Gmbh & Company KG, Germany), with extra bacterial lysis step. Before extraction, buccal swab was rubbed against the walls of 2 ml Eppendorf tube (Sarsted, Germany) containing 1 ml sterile PBS. After this, the buccal swab was cut into the tube. The tube was vortexed and incubated 1 hour at room temperature with shaking. The swab was dried and removed from the tube. After this, the tube was centrifuged 10 000 rpm for 10 minutes. Supernatant was removed and buffer T1 from the extraction kit was added with 25 µl Proteinase K and beads type B (Macherey-Nagel Gmbh & Company KG, Germany). Sample tubes were agitated for 3 minutes on a BulletBlender (NextAdvance, USA) at level 10. The samples were centrifuged 13 000 rpm for 3 minutes and incubated 20 minutes at 56°C. After this, standard protocol for human or animal tissue and cultured cells from NucleoSpin Tissue kit manual was followed from the step 3 (lyse sample) forward. After DNA extraction, DNA library was built using “16S metagenomics sequencing preparation instructions; preparing 16S ribosomal RNA gene amplicons for the Illumina MiSeq system”. Bacterial DNA amplification was done with PCR (Gulnaz Javan et al. Human Thanatomicrobiome Succession and Time Since Death, 2016) and the product was purified with AMPure XP-magnetic beads (Beckman Coulter, USA). Index PCR for the product was done with Illumina’s Nextera XT Index kit V2 set A (Illumina, USA) following the kit’s instructions. The size and purity of the libraries were confirmed with Fragment Analyzer (Agilent Technologies, USA). Library concentration was measured using Qubit 3 Fluorometer (Invitrogen, USA) and diluted to 4 nM. After library preparation, and sequencing was performed with MiSeq (Illumina, USA).

**Figure 1.**
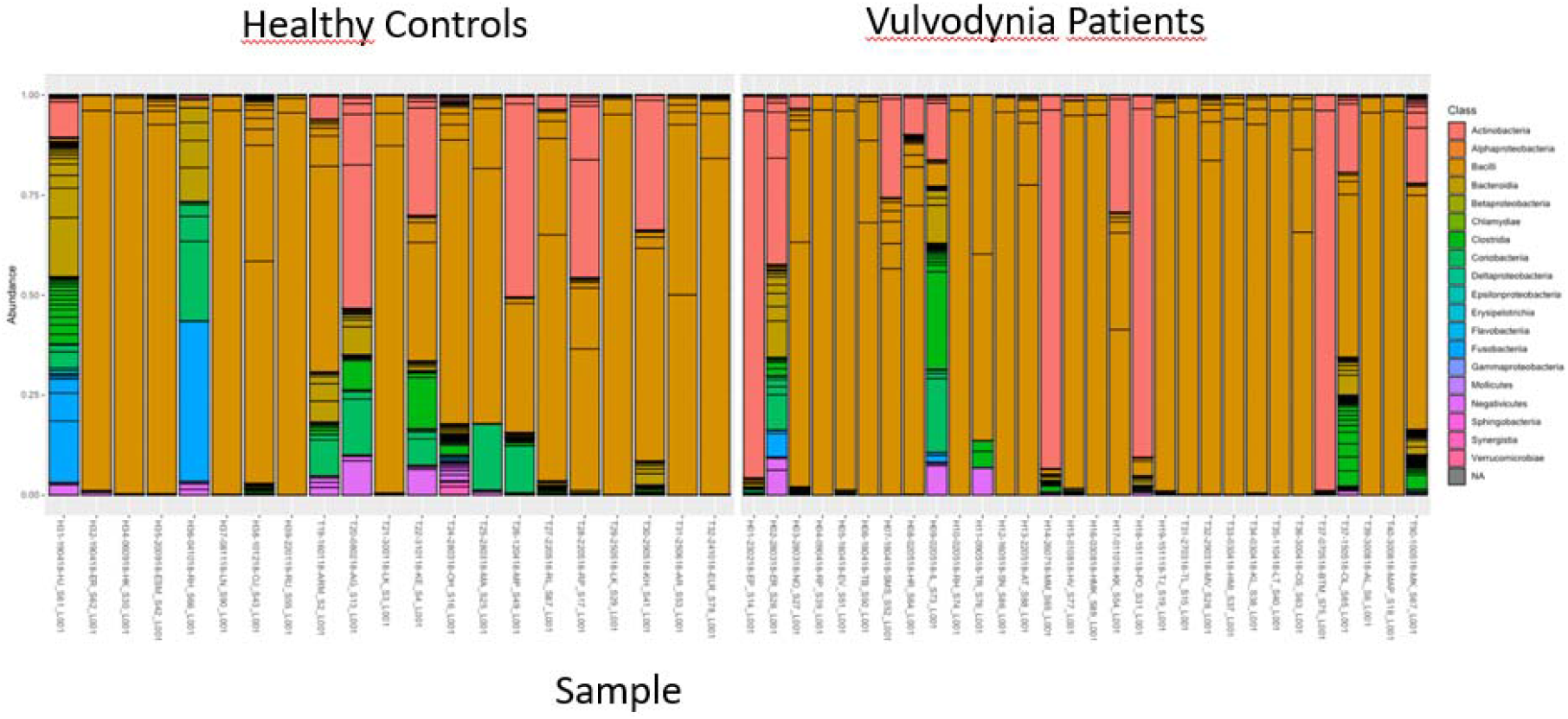
Stacked bar plot indicating the bacterial composition of vaginal samples from controls and vulvodynia patients.

The removal of primers from amplicon sequencing data was done using cutadapt tool^17^. Nucleotide bases with estimated probability of error less than 0.63 (default setting) were trimmed using filterAndTrim functionality implemented in DADA2 program^18^. Amplicon sequence variants (ASVs) were inferred using DADA2 approach. Chimers accounted for 5% of the sequences and were filtered out. Taxonomy assignment of the sequences were done using silva database (version 128)^19^. Bacterial contaminants were identified both manually and statistically using *decontam* R package ^20^ (See Supplementary text for the list of contaminants). Sequence data after contaminant filtration was used for downstream statistical analysis.

The statistical analyses were performed in R statistical software (version 3.5.3). Statistical difference between vulvodynia patients and controls was tested with permutational multivariate analysis of variance (PERMANOVA) test using Euclidean distance matrix implemented in *adonis* function of R package *vegan*. PERMANOVA test is based on assumption that there is homogeneity of dispersion within the compared groups. Validity of the assumption was tested by analyzing multivariate homogeneity of group dispersions using *betadisper* function in R package vegan. Differentially abundant taxa were determined using the DESeq2 package ^21^.

The statistical analysis comparing the demographic data on vulvodynia patients and controls was performed with Version 25 of IBM SPSS statistics software (IBM SPSS Statistics for Windows, Version 24.0. IBM Corp. 2016. Armonk, NY). Mann-Whitney U-test was used for statistical comparisons of different groups. A probability value of *P*□<□0.05 was considered as statistically significant.

## Results

The demographic data of all LPV patients, secondary LPV patients and controls are shown in Table 1. The patients had significantly lower BMI, were younger and more often nulliparous than controls (Table 1). Also the secondary vulvodynia patients were younger (p=0.001), had somewhat lower BMI (p=0.021) and were more often nulliparous (p=0.000) than controls. Ten out of 30 vulvodynia patients had an undefined day of the menstrual cycle (no bleeding/irregular bleeding) for following reasons: one was postmenopausal, and the rest (n=9) were using contraceptive method that prevents bleeding. Two control patients had an anatomic reason for not bleeding (Ashermann’s syndrome, cervical stricture), one control patient was in amennorrhea due to progestin only pill. The swab sample was taken in the median on the day 11 of the menstrual cycle (IQR 8.75-19.75) for both groups. Most of the patients had their sample taken on cycle day 15 or before (65.8%).

**Table 1.**
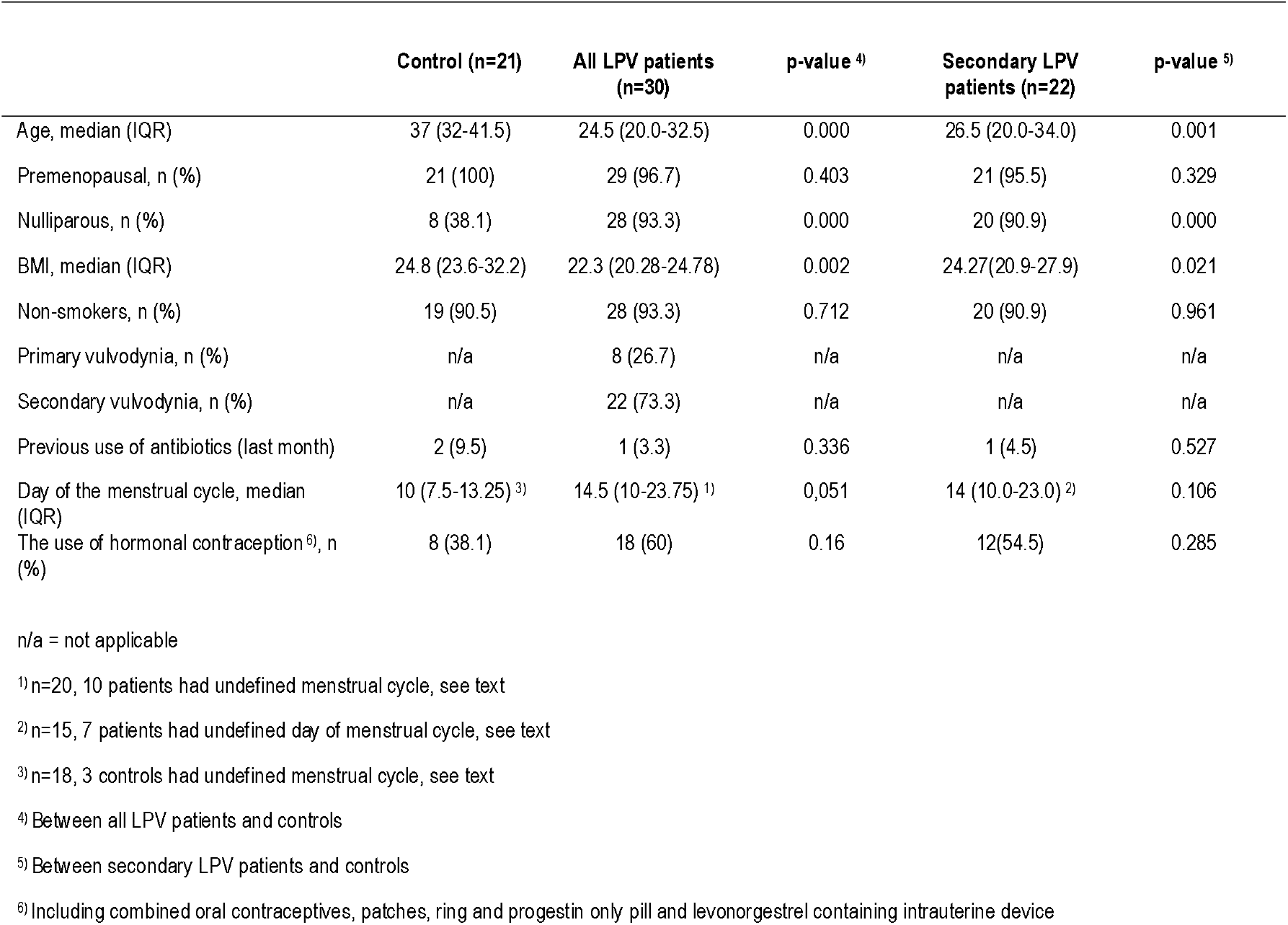
Demographic data of all LPV patients, secondary LPV patients and controls (Study IV)

**Table 2:**
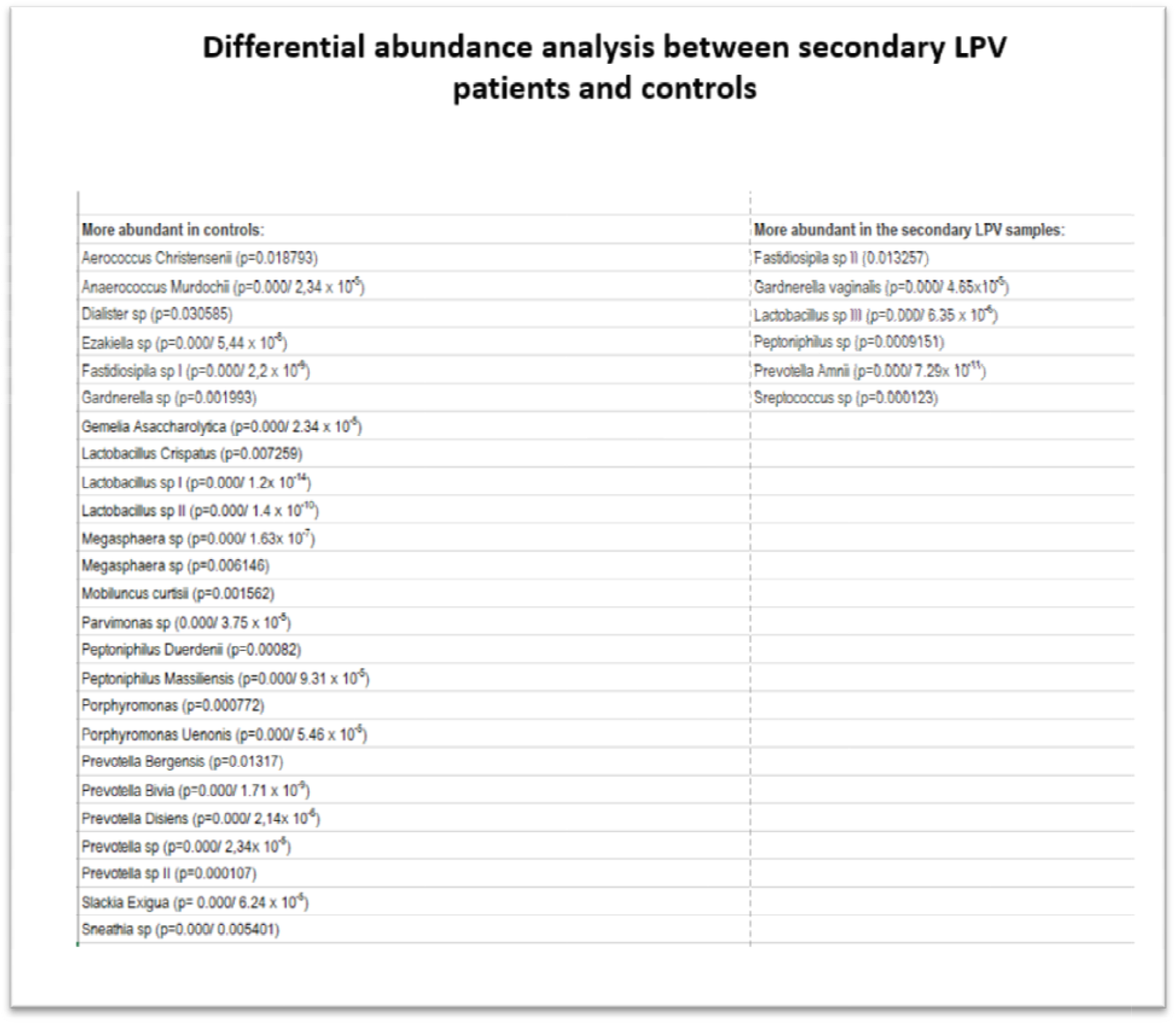
Differential abundance analysis results showing differences in bacterial abundances between secondary LPV patients and controls

### The microbiome of vulvar vestibulum in the study participants

Taxonomy analysis was performed in order to assess the overall bacterial composition in the vestibulum of all participants. Figure 1 shows the relative bacterial abundance in patients with primary or secondary vulvodynia at class level. *Bacillus* class (includes lactobacilli), was the most dominant taxa in majority of both control and patient samples (Figure 1). Other taxa contributing to the community were from classes *Actinobacteria, Clostridia, Fusobacteria* and *Negativicutes*. Distribution of different taxa seemed to be more representative among control individuals as compared to patients.

Alpha diversity estimates number of bacterial species (i.e. their amplicon sequence variant (ASV)) present in that microbiome (species richness) and evenness of their abundance Bacterial diversity in controls and LPV patients was assessed by three measures of alpha diversity: (a) **Chao1 index**, a richness estimator that represents the total number of distinct ASVs in a sample, (b) **Shannon index**, a proportion of taxa of one particular category (for example, species) divided by the number of total taxa found, and (c) **Simpson index**, which is quite similar to Shannon index, but gives more weight to dominant categories. This means that few rare categories with few taxa will not affect the diversity. Statistical difference in alpha diversities between controls and vulvodynia patients was tested with Wilcoxon rank sum test. Patients with secondary LPV showed reduced bacterial diversity compared to controls. A difference most closely approaching significance was found with Shannon index (p-value: 0.06), followed by Simpson (p-value: 0.08) and Chao1 (p-value: 0.14) indexes, respectively.

#### Beta diversity

Beta diversity compares the difference in bacterial composition between samples. It involves comparison of samples to each other by calculating the distance between each pair of samples. Here, Euclidean distance of all samples was calculated to all other samples. The beta diversity represented by the calculated distance matrix was summarized with principal coordinate analysis (PCoA) plot (Figure 3). PCoA plot helps in analyzing distances among samples by projecting the distances into two dimensional spaces. Each individual dot represents an individual sample, color-coded by disease status, with percent variance shown on each axis.

**Figure 2.**
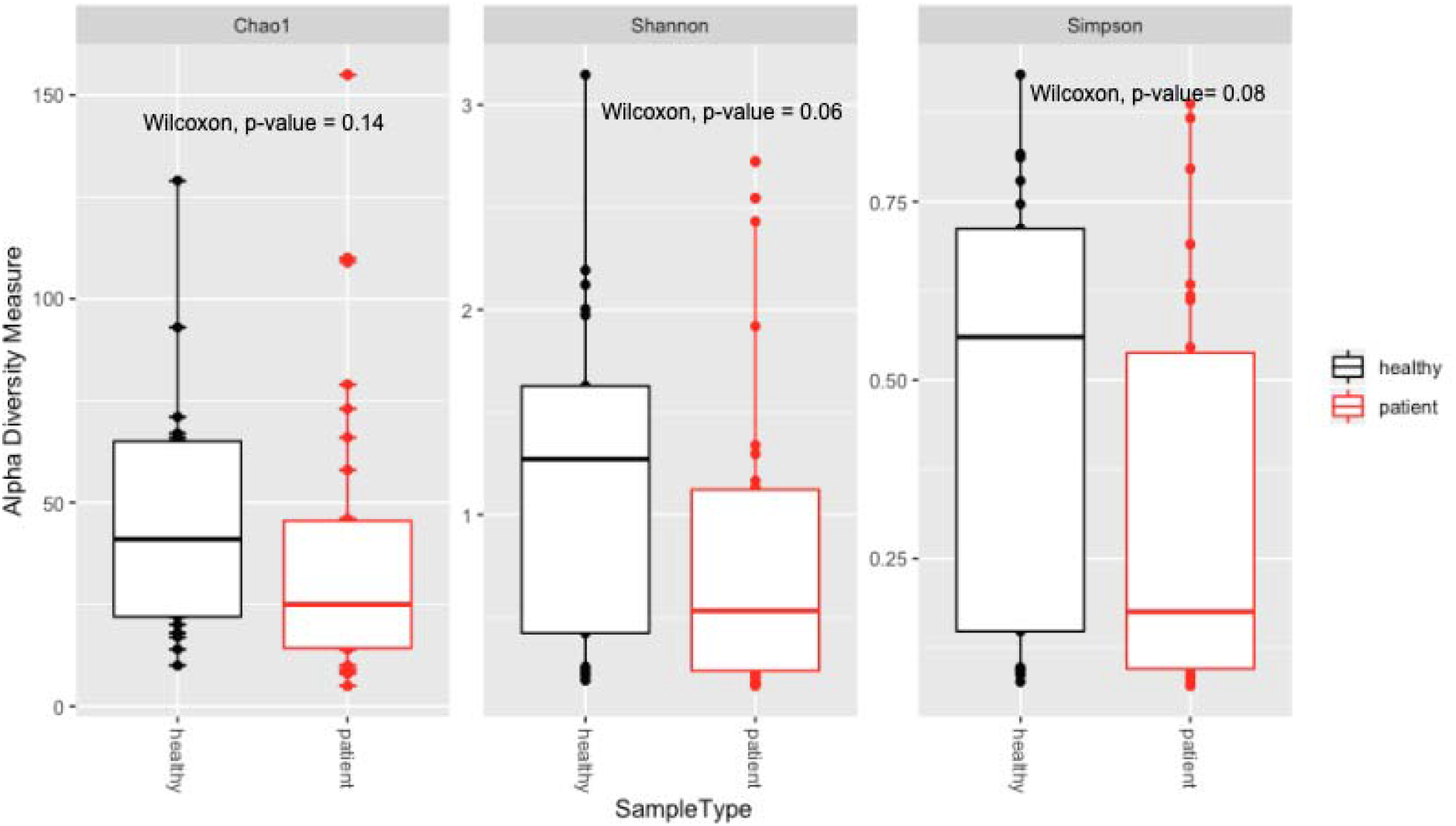
Boxplots comparing three measures of alpha diversity: Chao1, Shannon and Simpson (from left to right), between controls and LPV patients.

**Figure 3:**
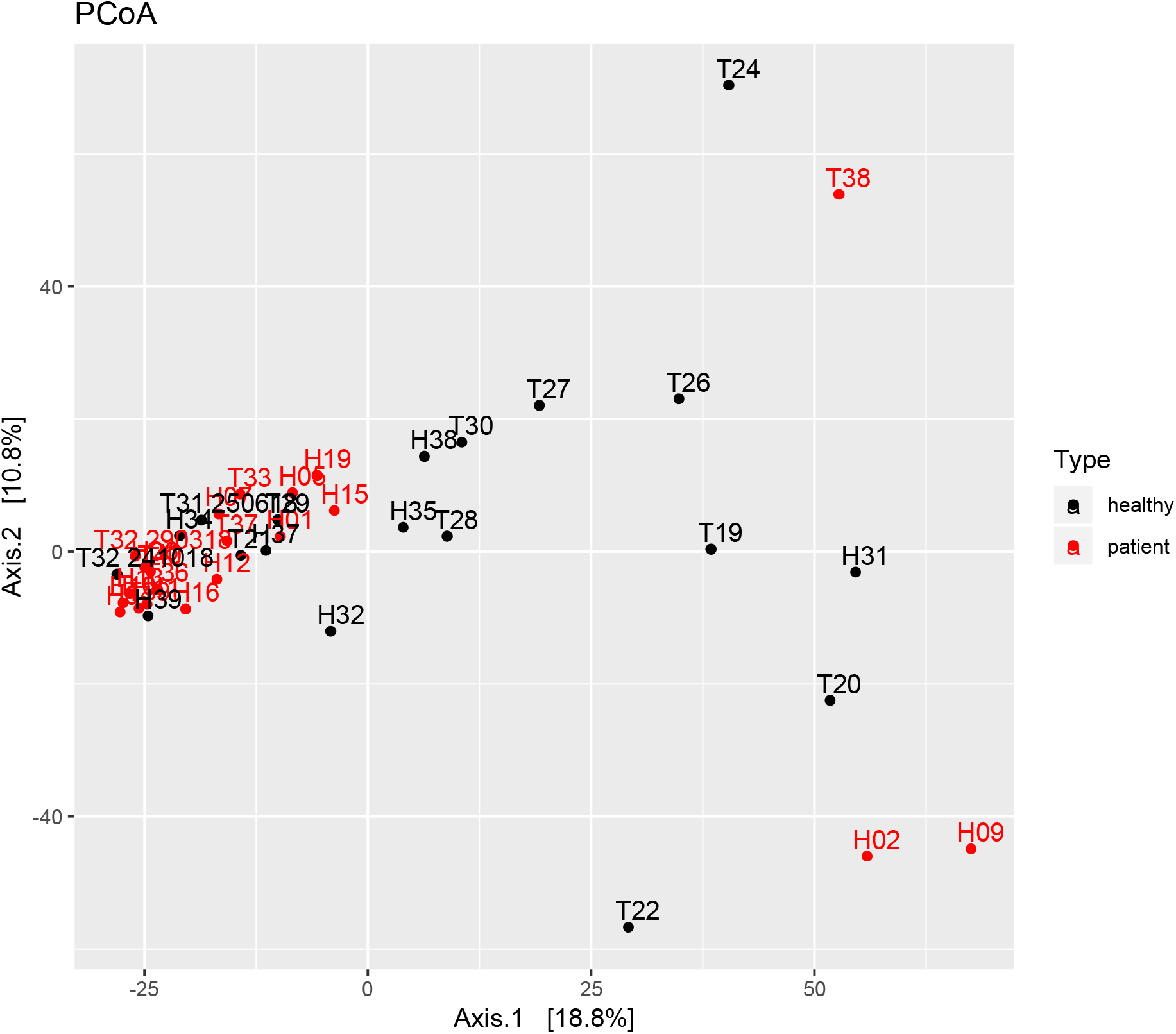
Principal coordinates analysis (PCoA) plot of the controls and secondary LPV patients, distinguished by black and red colors respectively.

Comparison of all LPV patients with healthy controls with adonis test did not show statistically significant difference. The test was repeated with only secondary LPV patients and controls. The idea was to maximize biological difference by excluding patients with primary condition which could have diluted the biological signal.

Significant statistical difference in bacterial composition between secondary LPV patients and healthy controls was seen (p= 0.05) explaining clustered vaginal microbiome communities seen in PCoA plot between controls and patients with secondary LPV.

Differences between bacterial composition of samples from healthy controls and secondary LPV patients were tested with two widely used statistical tests, betadisper and Adonis. Betadisper is used to test whether variance among the compared groups are homogenous. This must hold true in order to make reliable conclusions from Adonis test which tests whether or not the compared groups have similar bacterial compositions. A high p-value (p-value: 0.07) from betadisper test suggests that variances among the compared groups are not statistically different. This result allowed us to proceed with adonis test that suggests statistically significant difference (p= 0.05) between bacterial composition of controls and secondary vulvodynia patients. This difference was not significant when all LPV patients were compared to controls.

#### Differential abundance analysis

A statistically significant difference in bacterial composition of healthy controls and secondary LPV patients suggested by Adonis test motivated to investigate individual taxa responsible for the difference. This was achieved by differential abundance analysis using DESeq2 R program. Differential abundance analysis identified 31 bacterial taxa that contributed to the difference in bacterial composition between secondary cases and controls (p-value < 0.05). Bacteria taxa that were increased among patients were *Gardneralla vaginalis* (p<0.0001), *Peptoniphilus* sp (p<0.0001), *Prevotella amnii* (p<0.0001) and S*treptococcus* sp (p=0.0001). Several bacterial species were more common abundant among healthy controls reflecting reduced bacterial diversity among patients. The analysis identified 31 differentially abundant taxa.

## Discussion

We report here significant differences in total of 31 species in the vaginal microbiome between secondary vulvodynia patients and controls by using 16 s rRNA sequencing. To our knowledge, this is the first study to report significant differences between LPV patients and controls by using NGS.

Primary and secondary LPV have been suggested to have different pathomechanisms^5,22,23^. Goetsch et al found that primary LPV patients had increased nerve densities and milder degree of inflammation compared to secondary LPV in tissue samples analyzed with immunohistochemistry ^22^ (IHC). Secondary and primary LPV showed differences also in terms of neural hypertrophy, neural hyperplasia and the number of CD4-positive T-cells, which was more pronounced in the latter^23^. Also progesterone receptor density and estrogen receptor alpha expression was upregulated in primary LPV compared to secondary ^23^. Our study showed differences in 31 bacterial species collected from vulvar vestibulum suggesting that the initiation of secondary LPV could be the change in vulvar microbial environment caused by e.g. inflammation, which has been associated with the initiation of symptoms in previous studies^8,24,25^.

In the study of Ventolini et al, Lactobacillus crispatus was not found in samples from vulvodynia patients, assessed with quantitative PCR ^15^. This finding is in line with our finding that control samples had significantly more Lactobacillus crispatus. Likewise, Jayaram et al reported a tendency of L. Crispatus being the dominant bacteria more frequently in controls than LPV patients ^14^. They also found that Lactobacillus iners was more frequently dominant bacteria in LPV patients than controls and that Lactobacillus gasseri was present only in samples from vulvodynia patients ^14^. Group B betahemolytic streptococcus has been associated with minimal erythema, superficial fissures and vulvar pain ^26^, which are typical features of LPV. In our study, one streptococcus species was significantly more abundant in LPV patients.

There is no consensus on the best analysis method in 16S rRNA based amplicon study. We choose DADA2 because it has higher resolution as it analyzes data at individual sequence levels called as amplicon sequence variants (ASVs). Traditional methods such as QIIME and mothur analyze cluster of sequences based on some similarity threshold such as 97%. The clusters are called as operational taxonomic units (OTUs). DADA2 has been shown to be more sensitive, specific, comprehensive and reproducible as compared to traditional OTU based methods^18^. Taxonomy assignment in DADA2 is based on Ribosomal Database Project (RDP) naive Bayesian classifier. The classification algorithm is implemented in other popular analysis pipelines such as qiime and mothur as well. The algorithm achieves better confidence only upto genus level.

The differences in vestibular bacterial community reported above become potentially understandable when considering central pathophysiological features of LPV: the sensitization of nociceptors^27^ and vulvar hyperinnervation^22,23,28^. It has been previously hypothesized, that alterations in the vaginal flora and an immunological response involving candida could contribute to the development of vulvodynia^15^ Ventolini et al^15^ found changes in cytokine levels between patients and controls that could hypothetically act as a mediators in senzitizing free nociceptor-endings. Prolonged neuronal firing sensitizes the dorsal horn of the medulla nociceptors and causes peripheral sensitization and allodynia, both of which are important mechanisms in LPV related pain. Better understanding and further research of these neuroimmunological mechanisms could ultimately lead to first-line targeted therapies for secondary LPV^29^. Our study supports the finding that primary and secondary vulvodynia are different entities, suggested to develop with different pathomechanism, because differences in microbiome were detected only between secondary vulvodynia patients and controls.

The secondary LPV and control group varied significantly in BMI, as the patients had somewhat lower BMI possibly due to older median age of controls. BMI higher than 30 has been shown to associate with Finegoldia and Corynebacterium dominant vulvar microbiota, whereas the vulvar microbiota of patients whose BMI was lower than 30, vulvar microbiota was dominated by Lactobacillus^30^. However, in our study, both study groups had median BMI <25, even though groups had slightly, but significantly different BMI. As LPV patients were significantly younger than controls and variations in microbiome are shown to occur depending on the age^31^, this difference is another fact that has to be considered when interpreting these results. However, this kind of differences are unevitable in clinical setting studies. The use of hormonal contraceptives and the day of the menstrual cycle can affect the microbial flora of the vagina^13^ and vestibulum, however, there was not significant difference between the groups in these variables. Patients with previous antibiotic medication (<1 month) were excluded from the final analysis to prevent bias linked to the use of antibiotics.

### Strength and limitations of the study

To our knowledge, this is one of the few studies to compare the microbiome of LPV patients to controls, and first study to report significant differences in several bacteria collected from vulvar vestibulum.

Small sample size is a clear limitation of our study and all results must be interpreted with caution. 16s rRNA sequencing is a constantly evolving method. Even though we have been careful to identify and rule out “the kitome” and all other possible sources of contamination in the study material, one can never totally rule out the possibility of losing clinically meaningful bacteria in the sample processing.

A cross-sectional study does not prove a causality between inflammation and pain. Moreover, due to the study setting and the constant changes in microbial environment, this study does not reflect the microbiome status in the beginning of the pain symptoms. It could also be argued that the changes in the microbiome are *secondary* to the development of symptoms.

## Conclusion

We report here a significant difference of 31 bacteria from vulvar vestibulum when comparing secondary LPV patients to controls. If confirmed in a larger study setting, this difference in vulvar microbiome could serve as a possible target for prevention and/or treatment of secondary LPV.

## Data Availability

The data is available upon request from the corresponding author

